# APOE Genotype and Statin Response: Evidence from the UK Biobank Baseline Assessment and Linked Mortality Data

**DOI:** 10.1101/2024.12.13.24318982

**Authors:** Innocent G. Asiimwe, Andrea L. Jorgensen, Munir Pirmohamed, Multimorbidity Mechanism and Therapeutic Research Collaborative

## Abstract

**Introduction:** *APOE* genotype may influence response to statin therapy. We examined the relationship between *APOE* genotype, statin use, lipid biomarkers and mortality using data from the UK Biobank.

**Methods:** UK Biobank baseline assessment data and linked mortality records (389,843–452,189 participants) were analysed. Linear regression and Cox proportional hazards models assessed associations between *APOE* genotype, statin use, and lipid biomarkers (Apolipoprotein A, Apolipoprotein B, HDL cholesterol [HDLC], LDL cholesterol [LDLC], Lipoprotein A, Total Cholesterol, Triglycerides) as well as mortality, adjusting for clinical and genetic covariates.

**Results:** Significant interactions between *APOE* genotype and statin use were observed for most lipid biomarkers at the Bonferroni-adjusted threshold (*P* < 0.007), including Apolipoprotein A (*P* = 0.0065), Apolipoprotein B (*P* < 2.00e-16), LDLC, Total Cholesterol, and Triglycerides (all *P* < 2.00e-16), and HDLC (*P* = 0.0001). Lipoprotein A was not significant (*P* = 0.104). Population-level trends did not always translate to individual outcomes; for example, statin-treated *ε4ε4* carriers showed significant LDLC reductions but their LDLC levels remained higher than those of untreated *ε2ε2* individuals. *APOE* genotype was significantly associated with all-cause death (trend *P* < 2.00e-16) and cardiovascular-related death (*P* = 1.55e-10). The *ε4ε4* genotype had the highest risk, with respective hazard ratios of 1.51 (95% CI: 1.41– 1.62) and 1.54 (1.33–1.77). However, the *APOE*:statin use interaction was not significant.

**Conclusion:** The *APOE* genotype influences lipid biomarker levels, with statin use associated with favourable changes across all genotypes. The magnitude of these changes depends on both the APOE genotype and baseline lipid levels.

## Introduction

Cardiovascular diseases (CVD) continue to be the leading cause of death globally, with ischaemic heart disease accounting for the highest number of age-standardized deaths in 2021, despite the COVID-19 pandemic.^1^ Statins, also known as 3-hydroxy-3-methylglutaryl coenzyme A (HMG-CoA) reductase inhibitors, have been shown in randomized controlled trials (RCTs) to reduce both overall and CVD-related mortality.^2^ As a result, they are widely used for the primary and secondary prevention of cardiovascular disease.

The Apolipoprotein E (*APOE*) gene encodes the Apo E protein and is essential for lipid metabolism. It is present in lipoproteins such as very-low-density lipoproteins (VLDL) and certain high-density lipoproteins (HDL), playing a significant role in both CVD risk and statin response.^3-5^ Mutations in two SNPs (rs429358 and rs7412) give rise to three main isoforms (*ε2, ε3, ε4*) which influence lipid clearance by interacting with low-density lipoprotein (LDL) receptors. The *ε3* isoform, found in over half of the population,^3, 6^ is considered the wild-type. The *ε2* isoform binds poorly to LDL receptors, leading to slower lipid clearance and lower cholesterol levels through increased HMG-CoA and LDL receptor synthesis.^7-9^ In contrast, the *ε4* isoform increases lipid clearance, resulting in higher cholesterol levels. As a result, the *ε4* isoform is linked to higher CVD risk, while *ε2* is associated with lower risk.^7, 8^ However, in rare cases, individuals homozygous for *ε2* may develop familial dysbetalipoproteinemia, a condition marked by impaired lipid metabolism and increased CVD risk.^3, 10^

We recently conducted a meta-analysis (52 studies) to evaluate the impact of *APOE* genotype and carrier status on statin response.^11^ Compared to *ε3* carriers, *ε2* carriers showed greater percentage reductions in LDL cholesterol (LDLC), with similar reductions in total cholesterol (TC) and triglycerides (TG). We found that *ε4* carriers experienced less pronounced effects from statins, with smaller reductions in LDLC, TC and TG. Despite the inclusion of a significant number of studies, our analysis faced several limitations, including considerable heterogeneity (likely due to variations in study design, participant characteristics, genotyping procedures, etc.), a limited number of studies focusing on clinical outcomes such as mortality, an inability to account for specific statin types and doses used in the included studies, a lack of standardized reporting for genotypic and phenotypic data, and the inclusion of studies of unknown quality.

To address the above limitations, one approach is to utilize large datasets such as the UK Biobank,^12, 13^ which provides standardized genotypes and clinical phenotypes at the individual level, along with linked mortality data. For example, Welsh and colleagues analysed 346,686 participants from the UK Biobank who had no history of cardiovascular disease and were not taking statins.^14^ Their findings indicated that non-fasting total cholesterol and HDLC measurements were sufficient to predict the risk of both fatal (n=1656) and nonfatal (n=4560) cardiovascular events over a median follow-up period of 8.9 years.

Building on this, our study aimed to investigate the relationship between *APOE* genotype, statin use, and outcomes such as changes in lipid levels, as well as both all-cause and cardiovascular-related mortality, using data from the UK Biobank’s baseline assessment and linked mortality records.

## Methods

The reporting of this study adheres to the REporting of studies Conducted using Observational Routinely collected health Data (RECORD) statement^15^ (Table S1) and the STrengthening the Reporting Of Pharmacogenetic Studies (STROPS) guideline^16^ (Table S2).

### Data Sources

We used data from the UK Biobank, a large, population-based, prospective cohort that recruited over 500,000 participants aged 37 to 73 between 2006 and 2010 across 22 assessment centres in England, Scotland, and Wales.^12, 13^ At baseline, the UK Biobank collected extensive phenotypic, health, lifestyle, and genome-wide genotypic information through physical measurements, questionnaires, biochemical assays, and multi-modal imaging. Mortality records linked to participants are also available. The UK Biobank received ethical approval from the North-West Multicentre Research Ethics Committee (approval number: 11/NW/0382), and all participants provided written informed consent before data collection. The analyses conducted for this study were approved by the UK Biobank (application number: 56653).

### Study Participants

We conducted analyses using data from the baseline assessment and linked mortality records. We included adult participants who met the following criteria: a) had genomic data, b) reported consistent genetic and self-identified sex, c) did not have sex chromosome aneuploidy, d) were non-outliers for heterozygosity or missing rate, and e) were not related to other included participants, as previously described.^17^ Individuals with the *ε2ε2* genotype are at risk of developing familial dysbetalipoproteinemia, a condition that disrupts lipid metabolism and increases CVD risk.^3, 10^ For the clinical outcomes, we conducted a sensitivity analysis by excluding individuals with dysbetalipoproteinemia. This condition was defined as *ε2ε2* homozygotes with total cholesterol levels ≥ 200 mg/dL (5.2 mmol/L) and triglycerides ≥ 175 mg/dL (2.0 mmol/L).^18, 19^ Excluding these individuals was preferred over adjusting for dysbetalipoproteinemia to avoid multicollinearity, as all individuals with the condition had the APOE *ε2ε2* genotype.

### Outcomes and Follow-up

We analysed seven lipid biomarkers measured at baseline, detailed in the UK Biobank category 17518 (https://biobank.ctsu.ox.ac.uk/ukb/label.cgi?id=17518), which includes the resources “Biochemistry assay quality procedures” and “Companion document for serum biomarker data.” The lipid biomarkers were Apolipoprotein A (Apo A, g/L), Apolipoprotein B (Apo B, g/L), HDL cholesterol (HDLC, mmol/L), LDL directly-measured cholesterol (LDLC, mmol/L), Lipoprotein A (Lipo(a), nmol/L), Total Cholesterol (TC, mmol/L), and Triglycerides (TG, mmol/L).

For clinical outcomes, we examined all-cause mortality and CVD-related deaths. CVD death was defined using International Classification of Diseases (ICD)-10 codes (I10–15, I44–51, I20–25, I61–73) in line with the European Systematic Coronary Risk Evaluation (SCORE) guidelines, as previously described.^14, 20^ Participants were followed from their baseline assessment until the occurrence of a clinical outcome or the censor date (31st December 2022).

### Predictors

The exposure variables were *APOE* genotype and self-reported statin use. The six *APOE* genotypes (*ε2ε2, ε2ε3, ε2ε4, ε3ε3, ε3ε4* and *ε4ε4*) were determined based on the SNPs rs429358 and rs7412.^21^

Genotyping, imputation, and quality control measures have been previously detailed.^12, 22^ Statin use was identified using the Anatomical Therapeutic Chemical (ATC) Classification System code ‘C10AA’ (HMG CoA reductase inhibitors). Participants taking statins were identified through a previously established dictionary that maps UK Biobank self-reported medications to the ATC Classification System.^23^

The covariates adjusted for in the baseline analyses included age (in years) at recruitment, sex, body mass index (BMI, kg/m^2^), smoking status, alcohol consumption, race, level of physical activity, the genotyping array used, the Townsend index (a measure of socioeconomic status), and the first ten principal components representing genetic ancestry (for measurement and coding details, see our previous paper).^17^ Comorbidities were also adjusted for including diabetes mellitus (based on self-reported diabetes, doctor-diagnosed diabetes or use of anti-diabetic drugs), hypertension (based on systolic and diastolic blood pressures, and the use of antihypertensives [renin-angiotensin-aldosterone system inhibitors, beta-blockers, calcium channel blockers, and diuretics]), and cardiovascular disease (based on self-reported atrial fibrillation, coronary heart disease, stroke/transient ischaemic attack, peripheral vascular disease, and heart failure or doctor-diagnosed heart attack, angina, or stroke).^14, 24^

### Power analysis

Since we used an available cohort, we focused on calculating the available statistical power instead of determining the necessary sample size. For the lipid biomarkers, we used the power_envir.calc.linear_outcome function from the R package genpwr^25^ to calculate power, utilizing the sample sizes and standard deviations of the seven outcomes available from UK Biobank participants (https://biobank.ctsu.ox.ac.uk/ukb/label.cgi?id=17518). Assuming a target coefficient of determination (*R*^*2*^) of 1% for genetic, environmental, and gene-environment interaction effects, an additive mode of inheritance, a minor allele frequency as low as 1%, and approximately 17% statin use,^26^ we found that any sample size above 10,000 would yield statistical power close to 100%. This was determined using a Bonferroni-corrected *P*-value for the seven biomarkers (*P* = 0.05/7), indicating that our study was well-powered for all lipid biomarker outcomes.

For the two clinical outcomes (all-cause mortality and cardiovascular-related mortality), we used the powerEpiInt function from the R package powerSurvEpi^27^ to calculate statistical power. This function requires a pilot dataset and binary versions of the interacting variables (statins and *APOE*). For each genotype contrast compared to the reference genotype (*ε3ε3*), we randomly selected 1,000 participants from the UK Biobank cohort. As shown in Figure S1, assuming a significance level of 0.05, a sample size of 449,404 was sufficient to detect effect sizes for all-cause mortality across all genotype comparisons at an 80% power threshold, with a minimum hazard ratio (HR) of 1.1 (an HR of at least 1.2 was required for the *ε2ε2* genotype). For cardiovascular-related deaths, higher HRs were necessary to achieve over 80% power: HR = 2.3 for *ε2ε2*, 1.3 for *ε2ε3*, 1.5 for *ε2ε4*, 1.2 for *ε3ε4*, and 1.5 for *ε4ε4*.

### Missing Data

Participants with missing outcome data were excluded from the corresponding analyses. Less than 1% participants were missing data on at least one of the analysed predictors, and these were also excluded from analysis.

### Statistical Analysis

For the lipid outcomes, which were non-transformed and approximately normally distributed,^14^ we used linear regression models. In contrast, for mortality outcomes, we used Cox proportional hazard models with competing risks (considering two terminal states),^28^ where applicable, such as treating all-cause death as a competing risk for cardiovascular-related death. Each model included a Statin:*APOE* interaction term and adjusted for the covariates in the ‘Predictors’ section. The effects were reported as regression coefficients for linear regression and hazard ratios (HRs) for Cox regression, accompanied by 95% confidence intervals (CIs). To assess the proportional hazards assumption for the Cox regression models, we used the coxph function from the R package survival.^29^ Additionally, we employed Type III ANOVA tests from the car package^30^ to generate trend p-values for *APOE* genotype. This trend p-value offers a single measure of significance for the overall effect of the *APOE* genotype variable, considering it at the covariate level rather than just at individual factor levels. Statistical significance was set at *P* = 0.05, with multiple testing of the lipid biomarkers corrected using the Bonferroni method. All analyses were conducted in R (version 4.4.1).^31^

### Patient and public involvement

Patients and their representatives did not participate in formulating the research question or selecting the outcomes for this analysis. However, they are actively engaged with the UK Biobank.

## Results

### Participants

As of 13^th^ October 2024, a total of 502,191 UK Biobank participants had not withdrawn consent. After excluding 14,602 participants with missing genetic sex or mismatched clinical and genetic sex, 470 participants with sex chromosome aneuploidy, 963 participants who were heterozygosity or missing rate outliers, 33,946 participants with greater than 3rd-degree relatedness to included participants, and 2,806 participants missing *APOE* or other covariate data, the final analysed cohort consisted of 449,404 participants (Figure 1). This cohort included 389,843 for Apolipoprotein A (ApoA), 426,121 for Apolipoprotein B (ApoB), 392,018 for HDL Cholesterol (HDLC), 427,489 for LDL Cholesterol (LDLC), 342,592 for Lipoprotein A (Lipo(a)), 428,291 for Total Cholesterol (TC), 427,951 for Triglycerides (TG), and 449,404 for Mortality outcomes.

**Figure 1.**
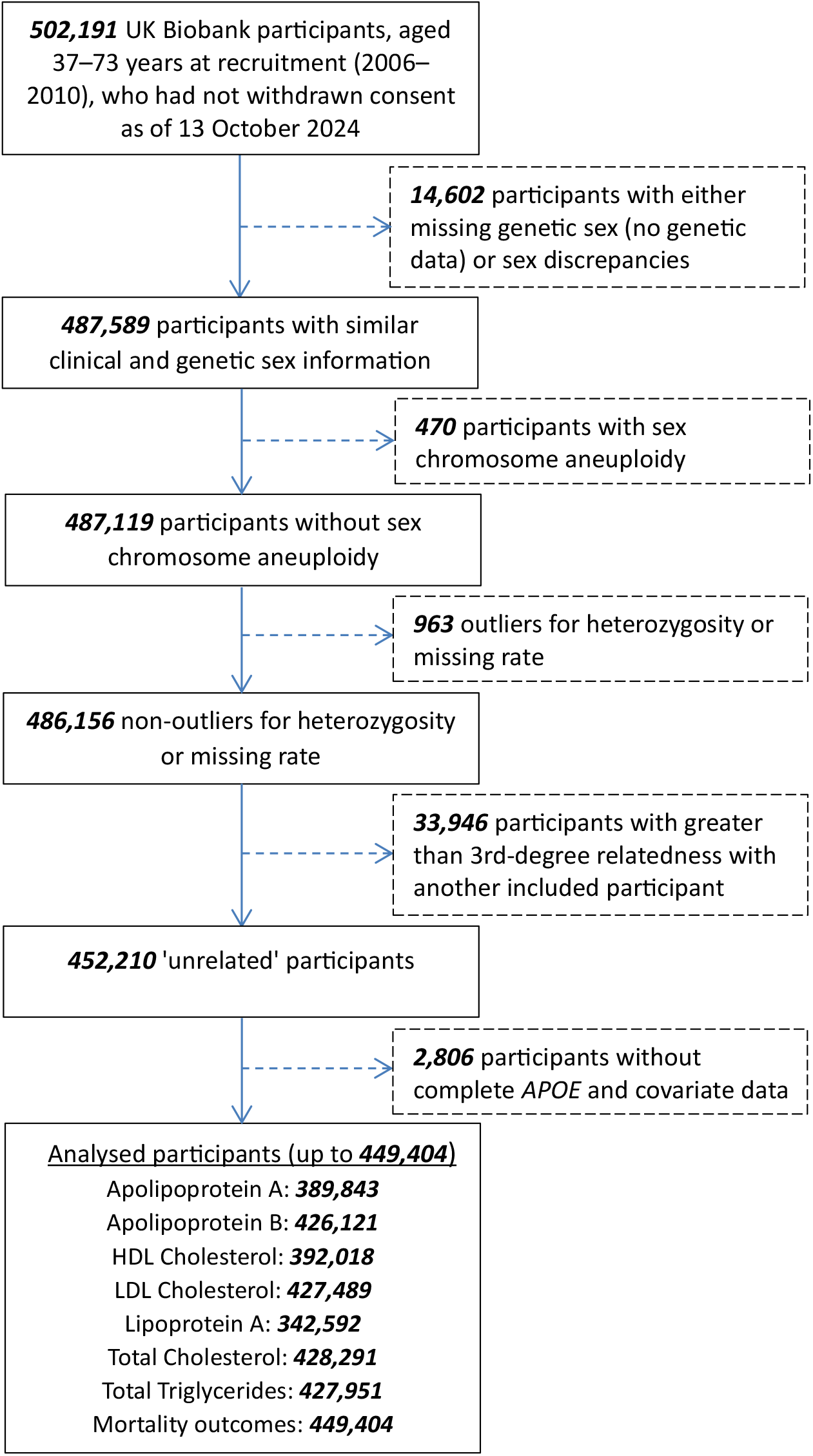
Flow chart for included participants. Bold values represent the total number of participants at each stage. *APOE* = Apolipoprotein E, HDL = high-density lipoprotein, LDL = low-density lipoprotein, MACE = major adverse cardiovascular events, VLDL = very low-density lipoprotein.

Table 1 presents the characteristics of these participants, categorized into those who were censored (*N* = 410,105; 91.3%) and those who died (*N* = 39,299; 8.7%). The mean age of participants was 56.6 years, with those who died being older (61.7 years). The cohort was predominantly female (53.9%) and White (94.2%). Body mass index (BMI) was slightly higher in those who died (28.3 kg/m^2^) compared to the overall mean of 27.4 kg/m^2^. The majority were non-smokers, with 54.5% never smoking, with current smoking being more prevalent among those who died (18.6% versus 9.7%). Alcohol use was common, with 91.8% reporting current use. Physical activity levels varied, with moderate activity being most frequent (59.6%). Antihypertensive medications (22.4%) and statins (16.5%) were common, particularly among those who died. Cardiovascular disease history (15.4%) and diabetes mellitus (5.3%) were more prevalent among those who died, who also had a shorter follow-up period (8.8 vs. 13.9 years).

**Table 1.**
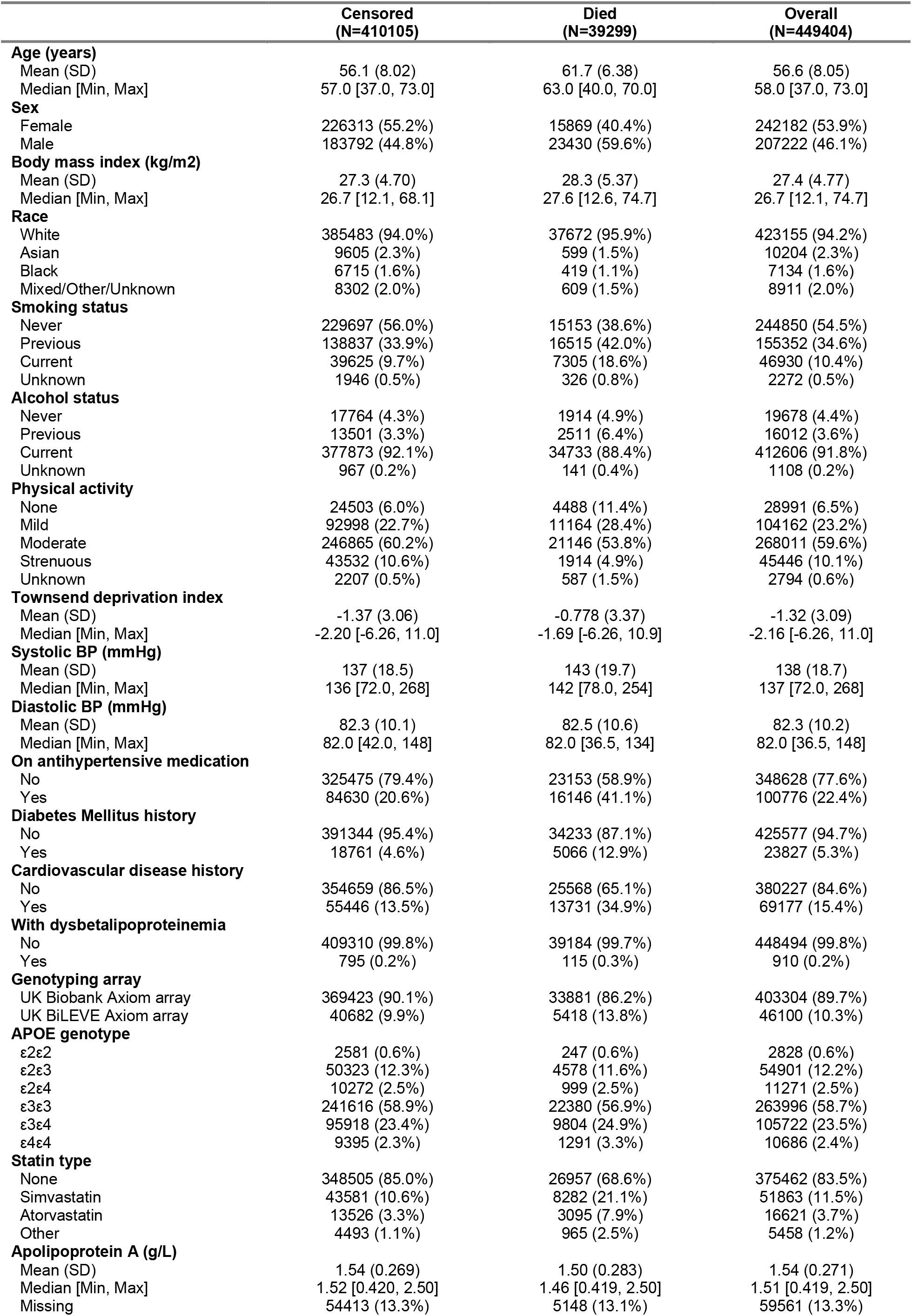
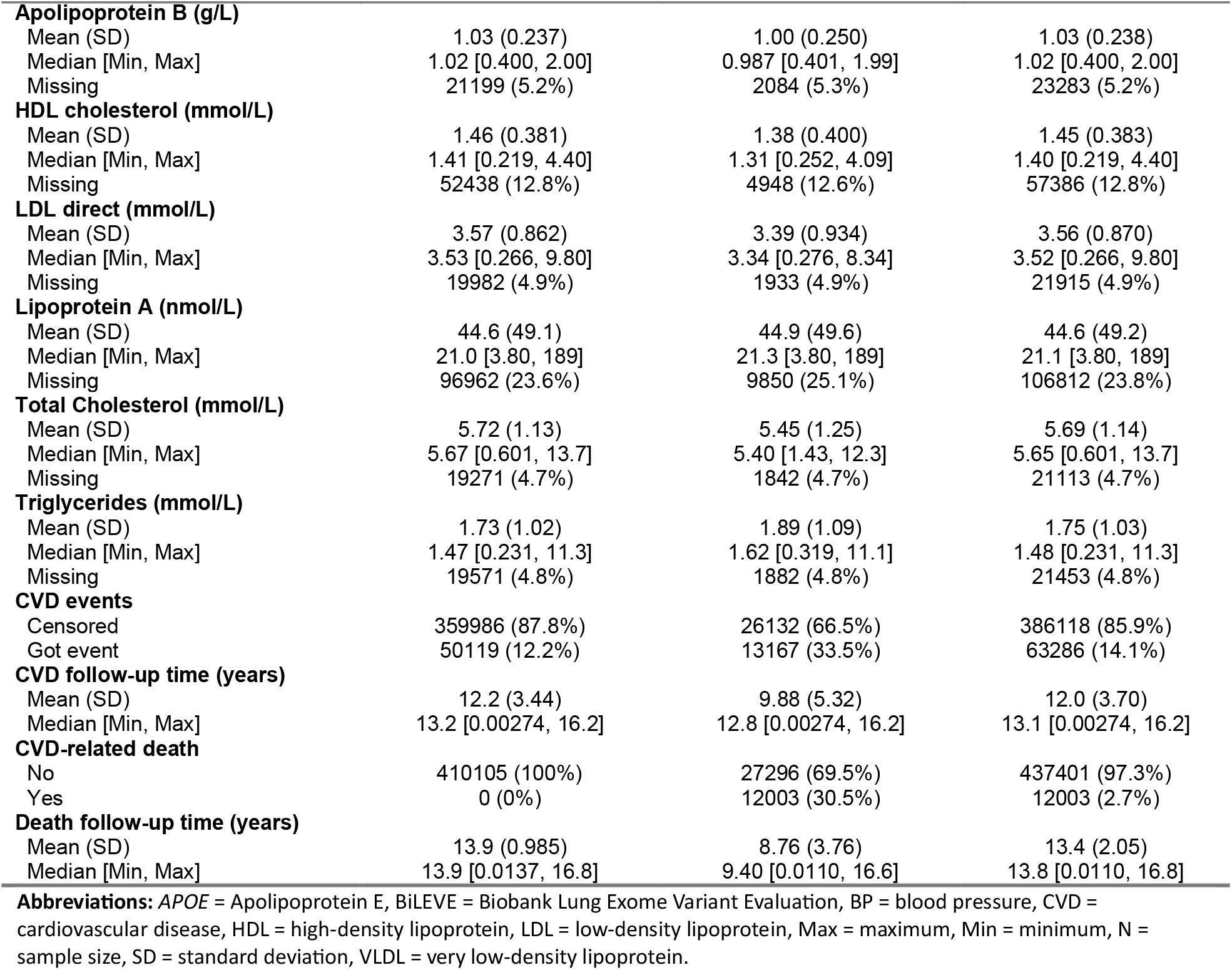
Characteristics of UK Biobank participants in the baseline assessment.

|  | **Censored**<br>**(N=410105)** | **Died**<br>**(N=39299)** | **Overall**<br>**(N=449404)** |
| --- | --- | --- | --- |
| **Age (years)** |  |  |  |
| Mean (SD) | 56.1 (8.02) | 61.7 (6.38) | 56.6 (8.05) |
| Median [Min, Max] | 57.0 [37.0, 73.0] | 63.0 [40.0, 70.0] | 58.0 [37.0, 73.0] |
| **Sex** |  |  |  |
| Female | 226313 (55.2%) | 15869 (40.4%) | 242182 (53.9%) |
| Male | 183792 (44.8%) | 23430 (59.6%) | 207222 (46.1%) |
| **Body mass index (kg/m2)** |  |  |  |
| Mean (SD) | 27.3 (4.70) | 28.3 (5.37) | 27.4 (4.77) |
| Median [Min, Max] | 26.7 [12.1, 68.1] | 27.6 [12.6, 74.7] | 26.7 [12.1, 74.7] |
| **Race** |  |  |  |
| White | 385483 (94.0%) | 37672 (95.9%) | 423155 (94.2%) |
| Asian | 9605 (2.3%) | 599 (1.5%) | 10204 (2.3%) |
| Black | 6715 (1.6%) | 419 (1.1%) | 7134 (1.6%) |
| Mixed/Other/Unknown | 8302 (2.0%) | 609 (1.5%) | 8911 (2.0%) |
| **Smoking status** |  |  |  |
| Never | 229697 (56.0%) | 15153 (38.6%) | 244850 (54.5%) |
| Previous | 138837 (33.9%) | 16515 (42.0%) | 155352 (34.6%) |
| Current | 39625 (9.7%) | 7305 (18.6%) | 46930 (10.4%) |
| Unknown | 1946 (0.5%) | 326 (0.8%) | 2272 (0.5%) |
| **Alcohol status** |  |  |  |
| Never | 17764 (4.3%) | 1914 (4.9%) | 19678 (4.4%) |
| Previous | 13501 (3.3%) | 2511 (6.4%) | 16012 (3.6%) |
| Current | 377873 (92.1%) | 34733 (88.4%) | 412606 (91.8%) |
| Unknown | 967 (0.2%) | 141 (0.4%) | 1108 (0.2%) |
| **Physical activity** |  |  |  |
| None | 24503 (6.0%) | 4488 (11.4%) | 28991 (6.5%) |
| Mild | 92998 (22.7%) | 11164 (28.4%) | 104162 (23.2%) |
| Moderate | 246865 (60.2%) | 21146 (53.8%) | 268011 (59.6%) |
| Strenuous | 43532 (10.6%) | 1914 (4.9%) | 45446 (10.1%) |
| Unknown | 2207 (0.5%) | 587 (1.5%) | 2794 (0.6%) |
| **Townsend deprivation index** |  |  |  |
| Mean (SD) | -1.37 (3.06) | -0.778 (3.37) | -1.32 (3.09) |
| Median [Min, Max] | -2.20 [-6.26, 11.0] | -1.69 [-6.26, 10.9] | -2.16 [-6.26, 11.0] |
| **Systolic BP (mmHg)** |  |  |  |
| Mean (SD) | 137 (18.5) | 143 (19.7) | 138 (18.7) |
| Median [Min, Max] | 136 [72.0, 268] | 142 [78.0, 254] | 137 [72.0, 268] |
| **Diastolic BP (mmHg)** |  |  |  |
| Mean (SD) | 82.3 (10.1) | 82.5 (10.6) | 82.3 (10.2) |
| Median [Min, Max] | 82.0 [42.0, 148] | 82.0 [36.5, 134] | 82.0 [36.5, 148] |
| **On antihypertensive medication** |  |  |  |
| No | 325475 (79.4%) | 23153 (58.9%) | 348628 (77.6%) |
| Yes | 84630 (20.6%) | 16146 (41.1%) | 100776 (22.4%) |
| **Diabetes Mellitus history** |  |  |  |
| No | 391344 (95.4%) | 34233 (87.1%) | 425577 (94.7%) |
| Yes | 18761 (4.6%) | 5066 (12.9%) | 23827 (5.3%) |
| **Cardiovascular disease history** |  |  |  |
| No | 354659 (86.5%) | 25568 (65.1%) | 380227 (84.6%) |
| Yes | 55446 (13.5%) | 13731 (34.9%) | 69177 (15.4%) |
| **With dysbetalipoproteinemia** |  |  |  |
| No | 409310 (99.8%) | 39184 (99.7%) | 448494 (99.8%) |
| Yes | 795 (0.2%) | 115 (0.3%) | 910 (0.2%) |
| **Genotyping array** |  |  |  |
| UK Biobank Axiom array | 369423 (90.1%) | 33881 (86.2%) | 403304 (89.7%) |
| UK BiLEVE Axiom array | 40682 (9.9%) | 5418 (13.8%) | 46100 (10.3%) |
| **APOE genotype** |  |  |  |
| ε2ε2 | 2581 (0.6%) | 247 (0.6%) | 2828 (0.6%) |
| ε2ε3 | 50323 (12.3%) | 4578 (11.6%) | 54901 (12.2%) |
| ε2ε4 | 10272 (2.5%) | 999 (2.5%) | 11271 (2.5%) |
| ε3ε3 | 241616 (58.9%) | 22380 (56.9%) | 263996 (58.7%) |
| ε3ε4 | 95918 (23.4%) | 9804 (24.9%) | 105722 (23.5%) |
| ε4ε4 | 9395 (2.3%) | 1291 (3.3%) | 10686 (2.4%) |
| **Statin type** |  |  |  |
| None | 348505 (85.0%) | 26957 (68.6%) | 375462 (83.5%) |
| Simvastatin | 43581 (10.6%) | 8282 (21.1%) | 51863 (11.5%) |
| Atorvastatin | 13526 (3.3%) | 3095 (7.9%) | 16621 (3.7%) |
| Other | 4493 (1.1%) | 965 (2.5%) | 5458 (1.2%) |
| **Apolipoprotein A (g/L)** |  |  |  |
| Mean (SD) | 1.54 (0.269) | 1.50 (0.283) | 1.54 (0.271) |
| Median [Min, Max] | 1.52 [0.420, 2.50] | 1.46 [0.419, 2.50] | 1.51 [0.419, 2.50] |
| Missing | 54413 (13.3%) | 5148 (13.1%) | 59561 (13.3%) |
| **Apolipoprotein B (g/L)** |  |  |  |
| Mean (SD) | 1.03 (0.237) | 1.00 (0.250) | 1.03 (0.238) |
| Median [Min, Max] | 1.02 [0.400, 2.00] | 0.987 [0.401, 1.99] | 1.02 [0.400, 2.00] |
| Missing | 21199 (5.2%) | 2084 (5.3%) | 23283 (5.2%) |
| **HDL cholesterol (mmol/L)** |  |  |  |
| Mean (SD) | 1.46 (0.381) | 1.38 (0.400) | 1.45 (0.383) |
| Median [Min, Max] | 1.41 [0.219, 4.40] | 1.31 [0.252, 4.09] | 1.40 [0.219, 4.40] |
| Missing | 52438 (12.8%) | 4948 (12.6%) | 57386 (12.8%) |
| **LDL direct (mmol/L)** |  |  |  |
| Mean (SD) | 3.57 (0.862) | 3.39 (0.934) | 3.56 (0.870) |
| Median [Min, Max] | 3.53 [0.266, 9.80] | 3.34 [0.276, 8.34] | 3.52 [0.266, 9.80] |
| Missing | 19982 (4.9%) | 1933 (4.9%) | 21915 (4.9%) |
| **Lipoprotein A (nmol/L)** |  |  |  |
| Mean (SD) | 44.6 (49.1) | 44.9 (49.6) | 44.6 (49.2) |
| Median [Min, Max] | 21.0 [3.80, 189] | 21.3 [3.80, 189] | 21.1 [3.80, 189] |
| Missing | 96962 (23.6%) | 9850 (25.1%) | 106812 (23.8%) |
| **Total Cholesterol (mmol/L)** |  |  |  |
| Mean (SD) | 5.72 (1.13) | 5.45 (1.25) | 5.69 (1.14) |
| Median [Min, Max] | 5.67 [0.601, 13.7] | 5.40 [1.43, 12.3] | 5.65 [0.601, 13.7] |
| Missing | 19271 (4.7%) | 1842 (4.7%) | 21113 (4.7%) |
| **Triglycerides (mmol/L)** |  |  |  |
| Mean (SD) | 1.73 (1.02) | 1.89 (1.09) | 1.75 (1.03) |
| Median [Min, Max] | 1.47 [0.231, 11.3] | 1.62 [0.319, 11.1] | 1.48 [0.231, 11.3] |
| Missing | 19571 (4.8%) | 1882 (4.8%) | 21453 (4.8%) |
| **CVD events** |  |  |  |
| Censored | 359986 (87.8%) | 26132 (66.5%) | 386118 (85.9%) |
| Got event | 50119 (12.2%) | 13167 (33.5%) | 63286 (14.1%) |
| **CVD follow-up time (years)** |  |  |  |
| Mean (SD) | 12.2 (3.44) | 9.88 (5.32) | 12.0 (3.70) |
| Median [Min, Max] | 13.2 [0.00274, 16.2] | 12.8 [0.00274, 16.2] | 13.1 [0.00274, 16.2] |
| **CVD-related death** |  |  |  |
| No | 410105 (100%) | 27296 (69.5%) | 437401 (97.3%) |
| Yes | 0 (0%) | 12003 (30.5%) | 12003 (2.7%) |
| **Death follow-up time (years)** |  |  |  |
| Mean (SD) | 13.9 (0.985) | 8.76 (3.76) | 13.4 (2.05) |
| Median [Min, Max] | 13.9 [0.0137, 16.8] | 9.40 [0.0110, 16.6] | 13.8 [0.0110, 16.8] |

**Abbreviations:** APOE = Apolipoprotein E, BiLEVE = Biobank Lung Exome Variant Evaluation, BP = blood pressure, CVD = cardiovascular disease, HDL = high-density lipoprotein, LDL = low-density lipoprotein, Max = maximum, Min = minimum, N = sample size, SD = standard deviation, VLDL = very low-density lipoprotein.

In terms of genetic data, *APOE* genotype distributions were as follows: *ε2ε2* (0.6%), *ε2ε3* (12.2%), *ε2ε4* (2.5%), *ε3ε3* (58.7%), *ε3ε4* (23.5%), and *ε4ε4* (2.4%). For lipid biomarkers, median levels for ApoA were 1.51 g/L, ApoB was 1.02 g/L, HDLC was 1.40 mmol/L, LDLC was 3.52 mmol/L, Lipo(a) was 21.1 nmol/L, TC was 5.65 mmol/L, and TG were 1.48 mmol/L. Missing values for these biomarkers ranged from 4.7% to 23.8%, with the highest missing rate for Lipo(a) (23.8%). Correlation analysis revealed strong associations between several lipid markers: ApoA and HDLC (Spearman’s correlation coefficient, *r* = 0.92), ApoB and LDLC (*r* = 0.96), ApoB and TC (*r* = 0.89), and LDLC and TC (*r* = 0.95) (Figure S2). Table S3 shows the same participant characteristics, along with additional details on the principal components of genetic ancestry, while Table S4 shows these characteristics stratified by each lipid biomarker.

### Lipid outcomes

Table S5 presents the associations between all analysed covariates and lipid biomarkers in the UK Biobank baseline analysis. Statin use and *APOE* genotype showed significant associations with all lipid biomarkers, with the weakest association for statin use having a *p*-value of 5.97e-10, and for *APOE* genotype, all p-values were <2.00e-16. Except for Lipo(a) (*P* = 0.104), all other biomarkers met the Bonferroni-adjusted significance threshold of *P* < 0.007 for the interaction between *APOE* genotype and statin use (ApoA *P* = 0.0065, ApoB *P* < 2.00e-16, HDLC *P* = 0.0001, LDLC *P* < 2.00e-16, TC *P* < 2.00e-16, and TG *P* < 2.00e-16).

Figure 2 visually illustrates these interactions by showing the median adjusted values of lipid biomarkers (second row), net changes (third row), percent changes (fourth row), and changes relative to the *ε3ε3* genotype (bottom row) across *APOE* genotypes. Statin users generally showed more favourable lipid profiles, particularly with reduced LDLC and TC levels. Compared to the *ε3ε3* genotype, carriers of the *ε4* allele experienced greater reductions in ApoB, LDLC, TC, and TG, as well as greater increases in ApoA and HDLC, indicating a larger benefit from statins. In contrast, those with the *ε2* allele demonstrated the opposite effects. However, these findings reflect population-level trends and may not translate directly to individual outcomes. For example, although statin-treated individuals with the *ε4ε4* genotype experienced the largest reductions in LDLC levels, the top row of Figure 2 reveals that statin-treated *ε4ε4* individuals still had higher LDLC levels than *ε2ε2* individuals who were not taking statins.

**Figure 2.**
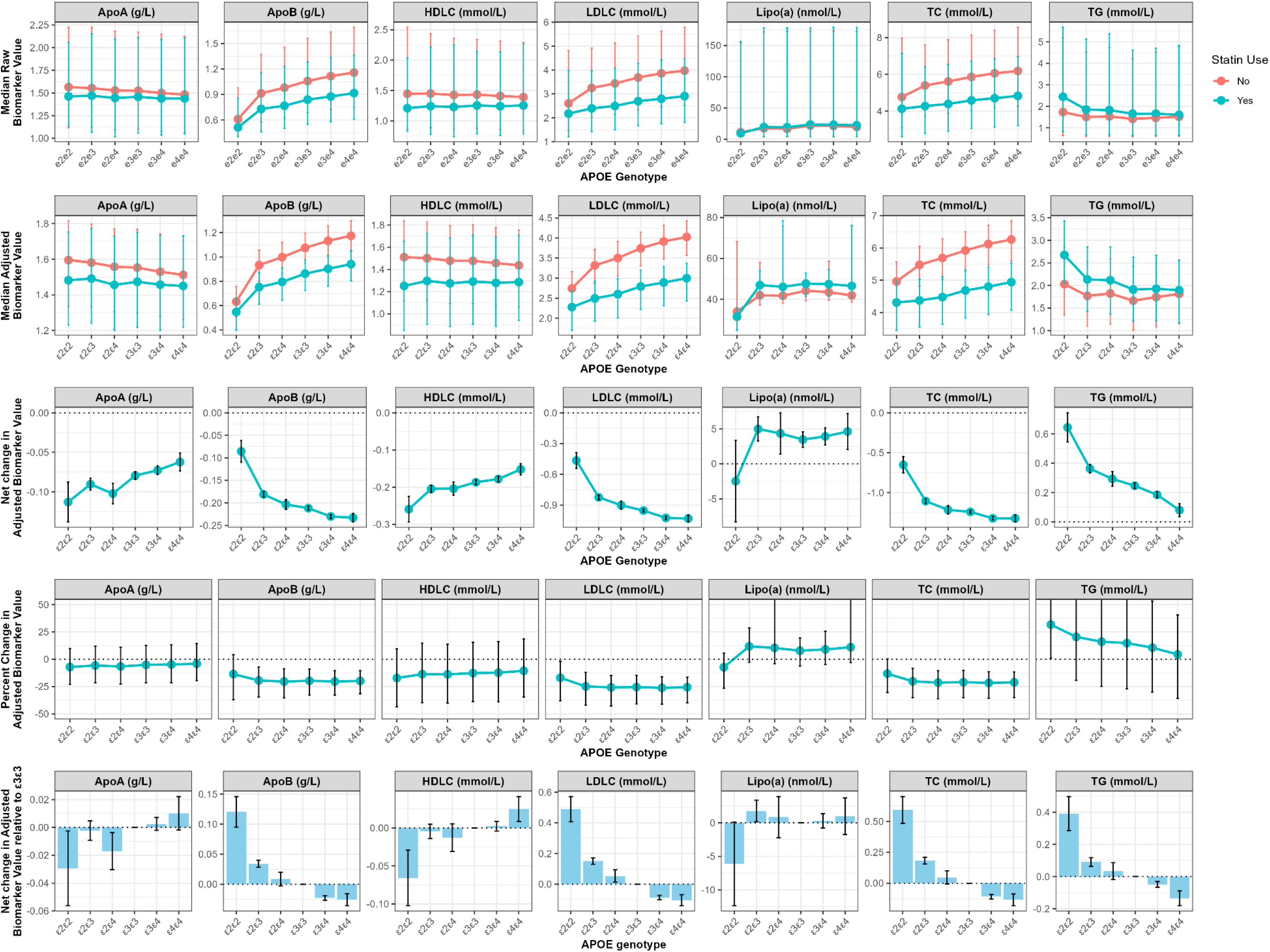
Associations between *APOE* genotype, statin use, and lipid biomarkers in the UK Biobank Baseline Analysis. This figure shows the median raw lipid biomarkers (top row), adjusted lipid biomarkers (second row), adjusted net changes (third row), adjusted percent changes (fourth row), and adjusted net change relative to the *ε3ε3* genotype (bottom row) across different *APOE* genotypes. The lipid biomarkers analysed include Apolipoprotein A (ApoA), Apolipoprotein B (ApoB), HDL cholesterol (HDLC), LDL cholesterol (LDLC), lipoprotein A (Lipo(a)), total cholesterol (TC), and triglycerides (TG). In the bottom row, for ApoA and HDLC (increase beneficial), positive values indicate more benefit with statins relative to the ε3ε3 genotype, while for other biomarkers (reduction beneficial), negative values indicate more benefit. Units for each biomarker are specified in their respective plots, with error bars representing 95% confidence intervals. Adjusted biomarker values are based on predictions from models adjusted for age at recruitment, sex, body mass index, smoking status, alcohol consumption, race, physical activity level, genotyping array, Townsend index, the first ten principal components of genetic ancestry, and the interaction between *APOE* genotype and statin use.

### Mortality outcomes

Table S6 summarizes the associations between all covariates and mortality outcomes, while Figure 3 highlights the impact of *APOE* genotype and statin use. In Panel A, the analysis of all-cause mortality shows that statin use significantly reduced the risk of death (hazard ratio [HR]: 0.92, 95% CI: 0.89–0.95, *P* = 2.37e-07). Stratification by *APOE* genotype revealed that individuals with the *ε4* allele, particularly *ε4ε4* carriers, had an increased risk of all-cause death (HR: 1.51, 95% CI: 1.41–1.62, *P* < 2.00e-16). In contrast, carriers of the *ε2* allele (*ε2ε2* and *ε2ε3*) showed a slightly lower or neutral risk. The interaction between statin use and APOE genotype was not statistically significant, suggesting that the mortality benefit of statins was generally consistent across genotypes.

**Figure 3.**
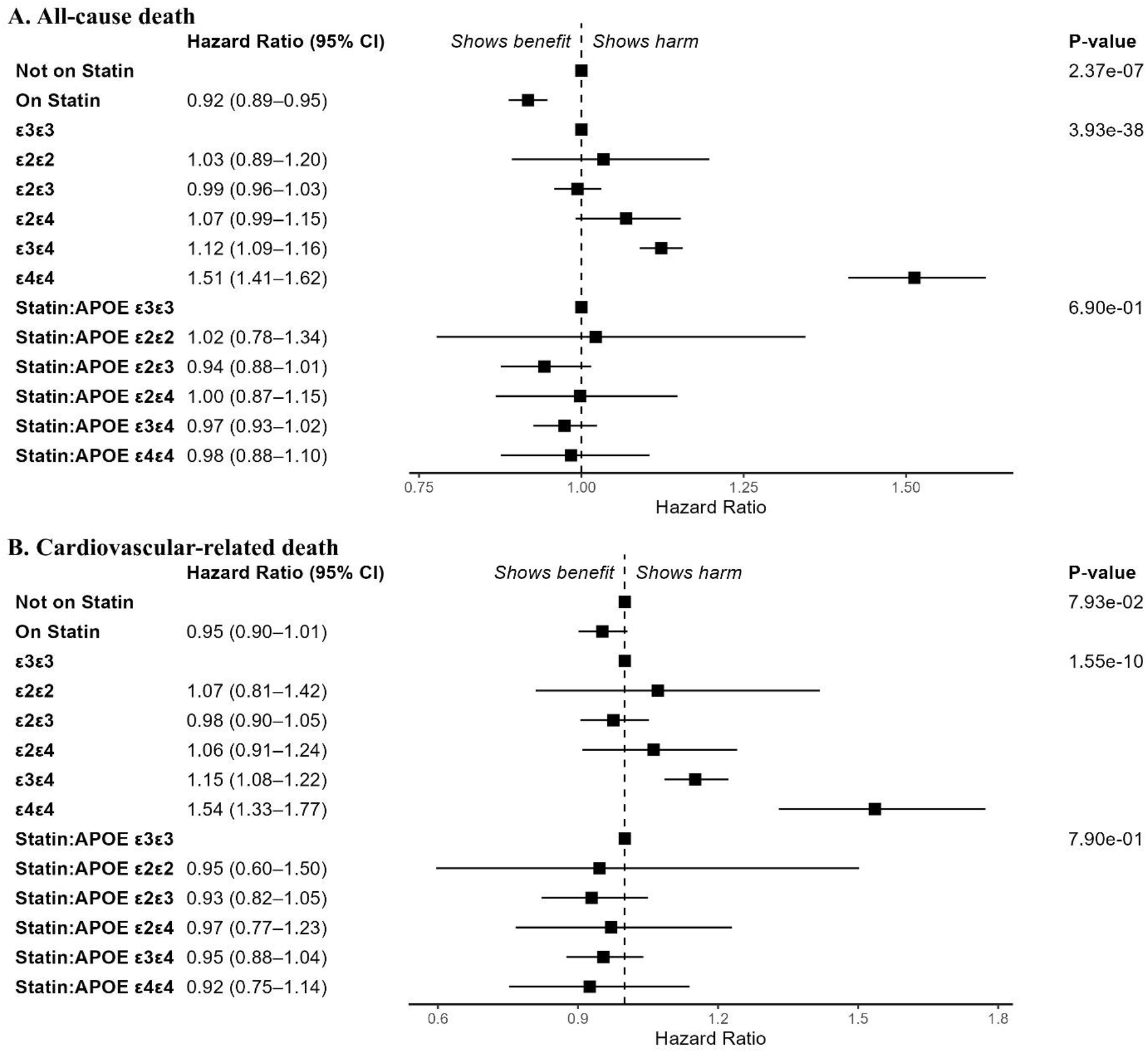
Associations between APOE genotype, statin use, and mortality outcomes in the UK Biobank. This figure presents hazard ratios and 95% confidence intervals (CIs) for (A) all-cause death and (B) cardiovascular-related death, stratified by statin use, *APOE* genotype, and the statin:*APOE* interaction. The analysis adjusts for multiple covariates, including age, sex, body mass index, race, smoking status, alcohol consumption, physical activity, Townsend deprivation index, systolic and diastolic blood pressure, use of antihypertensive medications, diabetes mellitus history, cardiovascular disease history, genotyping array, and the first ten principal components of genetic ancestry. *APOE* = Apolipoprotein E.

Sensitivity analyses (Table S6), which excluded 910 participants with dysbetalipoproteinemia, showed consistent results, reflecting the low proportion of excluded participants (0.2% of the total analysed). The results of the proportional hazards assumption test are presented in Figure S3. Based on a *P*-value < 0.05 or observable trends in time-varying hazards, we performed a sensitivity analysis that stratified time for relevant covariates using cut-offs at 7 and 12 years. This allowed the hazard to vary between the start of follow-up and year 7, between years 7 and 12, and from year 12 to the end of follow-up. The results, shown in Table S7, were generally consistent with the primary analysis. For example, the hazard ratios for statin use were 0.90, 0.92, and 0.93 across the three periods, with an average hazard ratio of 0.92, which was similar to that seen in the primary analysis.

Panel B of Figure 3 displays hazard ratios for cardiovascular-related death. Statin use again showed a protective effect (HR: 0.95, 95% CI: 0.90–1.01), but this was not statistically significant (*P* = 0.079).

Individuals with the *ε4* allele, especially those with the *ε4ε4* genotype, were at a higher risk of cardiovascular-related death (HR: 1.54, 95% CI: 1.33–1.77, *P* = 4.83e-09). Similar to all-cause mortality, the statin:*APOE* interaction was not significant for cardiovascular-related death.

## Discussion

Using data from the UK Biobank, we report the relationships between *APOE* genotype, statin use, lipid biomarkers, and mortality outcomes. Statin use (16.5%) was associated with favourable lipid outcomes, particularly for LDL cholesterol (LDLC) and total cholesterol (TC). As detailed in the 2019 European Society of Cardiology (ESC) and European Atherosclerosis Society (EAS) Guidelines for the management of dyslipidaemias^32^ and the 2018 American Cholesterol Clinical Practice Guidelines,^33^ statins inhibit 3-hydroxy-3-methylglutaryl coenzyme A (HMG-CoA) reductase, which is the rate-limiting enzyme in cholesterol synthesis in the liver. Reduced intracellular cholesterol leads to upregulation of LDL receptors on hepatocytes, enhancing the uptake of LDL from the bloodstream and consequently lowering plasma concentrations of LDL and other ApoB-containing lipoproteins (ApoB is the key structural protein in LDL particles), including triglyceride-rich particles. The effect of statins on LDLC is both statin- and dose-dependent, with high-intensity therapy typically resulting in at least a 50% reduction, and moderate-intensity therapy achieving a 30–49% reduction.^32, 33^ Given that LDLC is the main contributor to total cholesterol (with a correlation coefficient of 0.95 in our analysed data), TC levels also decrease significantly with statin treatment. Triglyceride (TG) levels are reduced by a smaller margin, typically 10– 20%.^32, 34^ Unlike LDLC, ApoB, TC, and TG that decrease with statin treatment, HDL cholesterol (HDLC) levels (ApoA being a component of HDL) have been reported to increase by 1–10%,^32, 33, 35^ while Lipoprotein A levels are generally unaffected by statins.^36^

Visual inspection of Figure 2 (top panel) confirms the expected reductions in ApoB, LDLC, and TC with statin use, alongside similar Lipoprotein A levels. However, unexpected trends are observed in ApoA and HDLC (lower median values) and TG (higher values) among statin users. These discrepancies can be attributed to the wide and overlapping confidence intervals, as well as the limitations inherent in cross-sectional, population-level studies like this one. For instance, it is hard to determine whether HDLC levels decreased due to statin use or if individuals prescribed statins initially had very low HDLC, which remained unchanged due to the modest effect of statins on HDLC. These findings highlight the importance of exercising caution when interpreting results from cross-sectional studies like ours.

Response to statins is variable and influenced by both clinical factors, such as age and sex, and genetic factors. Although we adjusted for several variables (see Tables S5–S7), our primary focus was the genetic influence of the *APOE* genotype. The distribution of *APOE* alleles in our study was consistent with prior population studies,^3, 6^ where the wild-type (*ε3*) was found in approximately 60% of the participants. To obtain the genetic influence, we included interactions between *APOE* genotype and statin use in our analysis. Except for Lipoprotein A, significant interactions were observed between *APOE* and statin use for all other lipid biomarkers analysed. Since Lipoprotein A levels are generally unaffected by statins,^36^ the lack of interaction for Lipoprotein A suggests that the lack of statin effect is consistent across *APOE* genotypes, or that any effect was too small to detect.

Visual inspection of Figure 2 (bottom three panels) shows that, compared to the *ε3ε3* genotype, carriers of the *ε4* allele experienced greater reductions in ApoB, LDLC, TC, and TG, as well as greater increases in ApoA and HDLC. This may suggest a larger benefit from statins, which contrasts with previous studies, which reported that *ε2* carriers generally respond better to statins than those with the *ε3ε3* genotype, while *ε4* carriers showed a poorer response.^7, 9, 37-40^ The top panel of Figure 2 may help explain these results. Using LDLC as an example, it can be seen that non-statin *ε4* carriers have much higher LDLC levels compared to non-statin *ε2* carriers. As observed by Tavintharan et al.,^41^ if baseline (pre-treatment) LDLC levels are much higher in *ε4* carriers, these individuals may show greater percentage reductions in response to statin therapy compared to *ε2* or *ε3* carriers with lower baseline levels. Although Figure 2 shows that *ε4* carriers receive the greatest benefit from statins at a population level, it also indicates that *ε4* carriers are the most resistant to statin therapy, as statin-treated *ε4ε4* individuals still had higher LDLC levels compared to untreated *ε2ε2* individuals. To address some of the limitations of population-level analysis, we provide individual-level analyses in another paper,^42^ utilizing electronic health record data from the UK Biobank and the All of Us research program.

A 2005 meta-analysis of 14 randomized controlled trials (RCTs) involving 90,056 individuals reported a 19% reduction in coronary mortality (risk ratio [RR]: 0.81; 95% CI, 0.76–0.85) per 1 mmol/L (38.7 mg/dL reduction in LDLC over a five years.^43^ A 2020 meta-analysis of 53 RCTs (329,897 participants) found a 15% reduction in cardiovascular mortality risk (RR: 0.85; 95% CI, 0.81–0.89) per 1 mmol/L LDLC reduction, with significant effects only at baseline LDLC >100 mg/dL.^44^ Additionally, a prospective cohort study of 14,035 adults who participated in the National Health and Nutrition Examination Survey III (1988–1994) showed increased cardiovascular mortality risk with both very low (<70 mg/dL; HR: 1.60; 95% CI, 1.01–2.54) and very high LDLC (≥ 190 mg/dL; HR: 1.49; 95% CI, 1.09–2.02).^45^ Although our study could not analyse individual net or percentage LDLC changes, we observed that statin use was associated with reduced all-cause mortality (HR: 0.92; 95% CI, 0.89–0.95) and a non-significant reduction in cardiovascular mortality (HR: 0.95; 95% CI, 0.90–1.01). Consistent with known biological mechanisms,^7, 8^ higher mortality risks were observed in *ε4* carriers, particularly *ε3ε4* (all-cause HR: 1.12, 95% CI 1.09–1.16; cardiovascular HR: 1.15, 95% CI 1.08–1.22) and *ε4ε4* genotypes (all-cause HR: 1.51, 95% CI 1.41–1.62; cardiovascular HR: 1.54, 95% CI 1.33–1.77). However, the interaction between statin use and *APOE* genotype was not significant, suggesting no genotype-specific differences in statin effects on mortality or an undetectably small effect.

Our study had some limitations. First, the study population may not be representative of the general population, as UK Biobank participants tend to be healthier overall.^46^ The cross-sectional study design used for lipid biomarkers as outcomes did not capture temporal relationships, and population-level analyses were unable to reflect changes in biomarkers at the individual level. Additionally, available covariates were limited – we were unable to adjust for statin type, dose, or treatment adherence, as these data were not available in the baseline dataset. Unlike some studies,^11^ we did not report *APOE* carrier status. Instead, we focused on *APOE* genotype for two main reasons: (a) there is no consensus on how to handle the *ε2ε4* genotype – whether it should be excluded or classified as an *ε2, ε3*, or *ε4* carrier; and (b) grouping *ε2ε2* and *ε2ε3* as *ε2* carriers could be misleading, as the *ε2ε2* genotype is associated with an increased risk of familial dysbetalipoproteinemia.^3, 10, 11^ Despite these limitations, our study had several strengths, including the use of a large cohort and the testing of assumptions, such as the proportional hazards assumption and the impact of retaining participants with dysbetalipoproteinemia, through sensitivity analyses.

In conclusion, we investigated the relationship between *APOE* genotype, statin use, lipid biomarkers, and mortality using data from the UK Biobank. While *APOE* genotype may interact with statins to influence lipid biomarkers, the interaction between *APOE* genotype and statin use was not statistically significant. However, both statin use and *APOE* genotype independently influenced statin response, with individuals carrying the *ε3ε4* and *ε4ε4* genotypes showing increased mortality risks. These findings add to the existing evidence base and provide insights into personalized lipid management strategies using statins with the aim of improving clinical outcomes, including reducing mortality risk.

## Conflict of interest

M.P. currently receives partnership funding, paid to the University of Liverpool, for the following: MRC Clinical Pharmacology Training Scheme (co-funded by MRC and Roche, UCB, Eli Lilly and Novartis), and the MRC Medicines Development Fellowship Scheme (co-funded by MRC and GSK, AZ, Optum and Hammersmith Medicines Research). He has developed an HLA genotyping panel with MC Diagnostics but does not benefit financially from this. He is part of the IMI Consortium ARDAT (www.ardat.org); none of these of funding sources have been used for the current research. All other authors declared no competing interests for this work.

## Funding

This work was supported by the Medical Research Council [MR/V033867/1; Multimorbidity Mechanism and Therapeutics Research Collaborative].

## Data Availability

The data that support the findings of this study are available from UK Biobank (https://www.ukbiobank.ac.uk/) with the permission of UK Biobank.

## Acknowledgements

This research has been conducted using the UK Biobank Resource under Application Number 56653.

## Notes

### Multimorbidity Mechanism and Therapeutic Research Collaborative

Adam Butterworth, Alasdair Warwick, Alba Fernandez-Sanles, Albert Henry, Alvina G Lai, Amanda Roberts, Andrea Jorgensen, Ana Torralbo, Anoop D Shah, Aroon Hingorani, Arturo Gonzalez-Izquierdo, Maria Carolina Borges, Caroline Dale, Chris Finan, Claudia Langenberg, Daniel C Alexander, Deborah Lawlor, Diana Dunca, Eda B Ozyigit, Amand F Schmidt, Harry Hemingway, Honghan Wu, Innocent G Asiimwe, Jasmine Gratton, Jean Gallagher, Jorgen E Engmann, Lauren E Walker, Victoria L Wright, Magdalena Zwierzyna, Margaret Ogden, Martin Cox, Mary Mancini, Michail Katsoulis, Mira Hidajat, Munir Pirmohamed, Natalie Fitzpatrick, Nishi Chaturvedi, Rashmi Kumar, Rohan Takhar, Sandesh Chopade, Simon Ball, Spiros Denaxas, Tina Shah, Valerie Kuan, Nikita Hukerikar, Reecha Sofat, Frances Bennett, David Ryan, Maik Pietzner.

## Notes

### Author Declarations

The UK Biobank received ethical approval from the North-West Multicentre Research Ethics Committee (approval number: 11/NW/0382), and all participants provided written informed consent before data collection. The analyses conducted for this study were approved by the UK Biobank (application number: 56653).

