## Supplementary Figures for "APOE Genotype and Statin Response: Evidence from the UK Biobank Baseline Assessment and Linked Mortality Data"

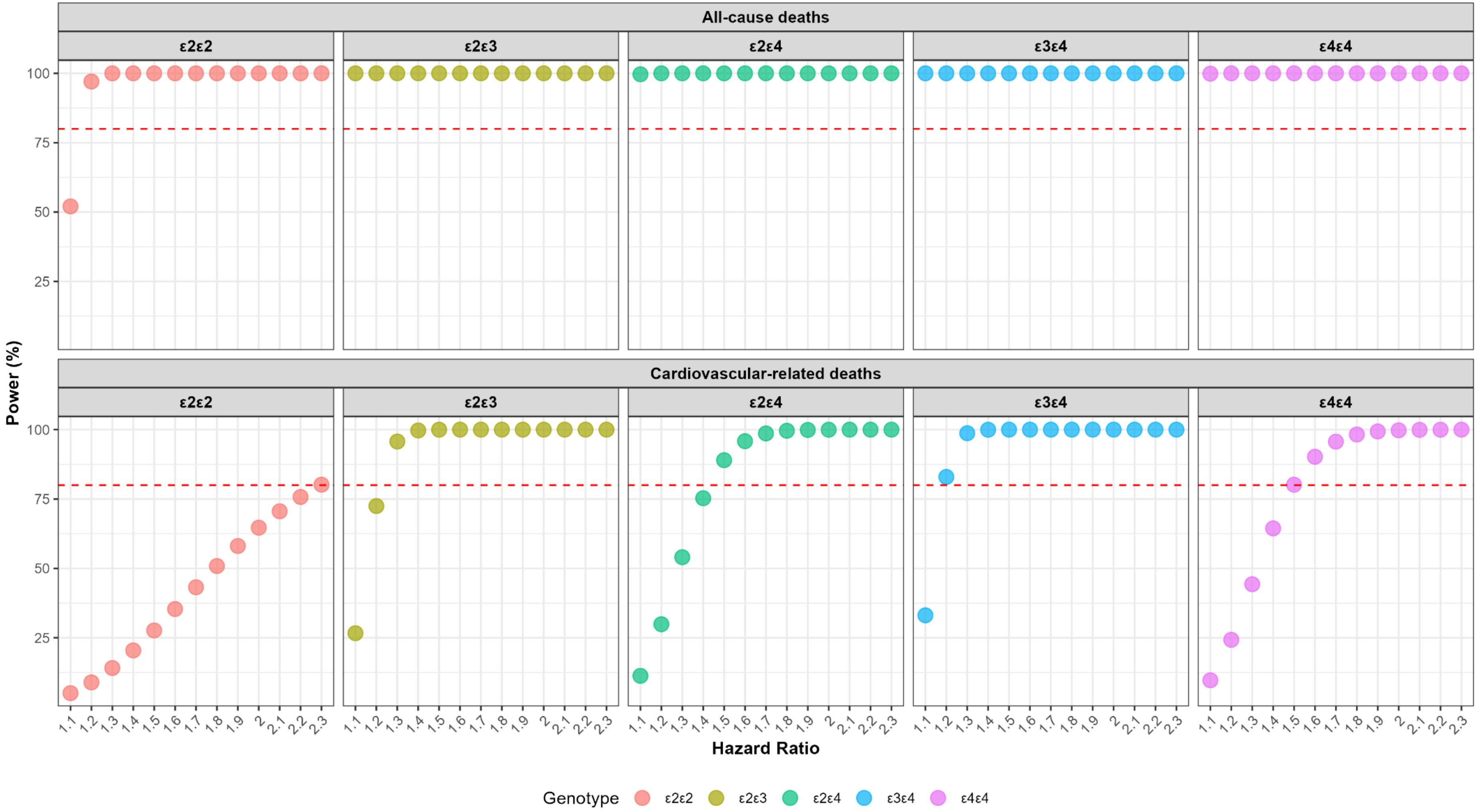


**Figure S1. Power analysis for varying hazard ratios for each *APOE* genotype, assuming a sample size of 449,404, with *ε3ε3* as the reference genotype.** Panels display power (%) to detect hazard ratios for all-cause deaths (top) and cardiovascular-related deaths (bottom) across the genotypes *ε2ε2*, *ε2ε3*, *ε2ε4*, *ε3ε4*, and *ε4ε4*. The dashed red line indicates the 80% power threshold.

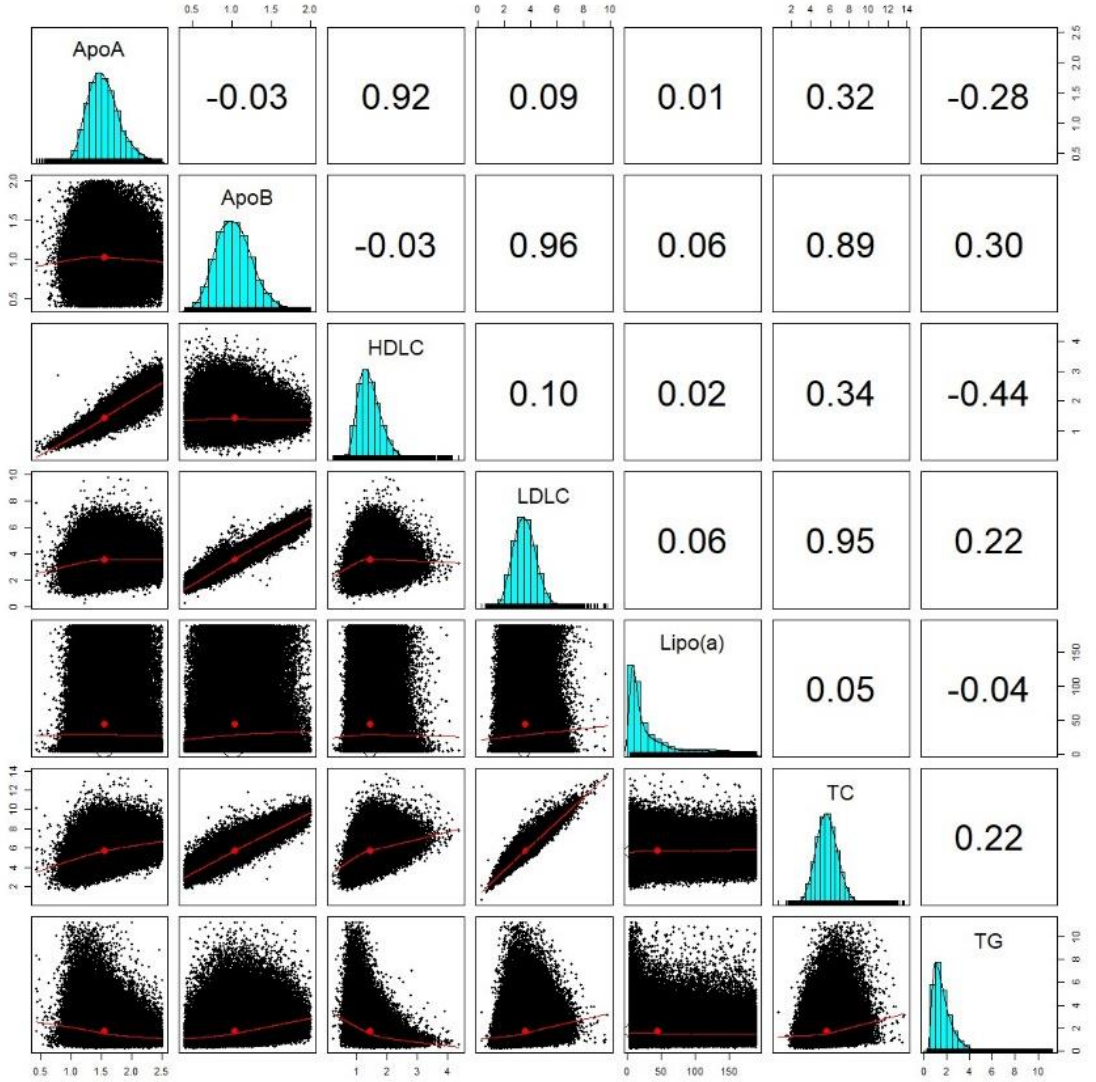


**Figure S2. An enhanced scatterplot matrix showing correlations between the lipid biomarkers in the UK Biobank Baseline Analysis (16.5% on statins).** This figure presents a correlation matrix and pairwise scatter plots for key lipid biomarkers: Apolipoprotein A (ApoA), Apolipoprotein B (ApoB), HDL cholesterol (HDLC), LDL cholesterol (LDLC), lipoprotein A (Lipo(a)), total cholesterol (TC), and triglycerides (TG). The upper triangles show Pearson correlation coefficients between the biomarkers, the diagonals display the distribution of each biomarker, and the lower triangles shows scatter plots with locally weighted smoothing (red lines) to depict relationships between the variables.

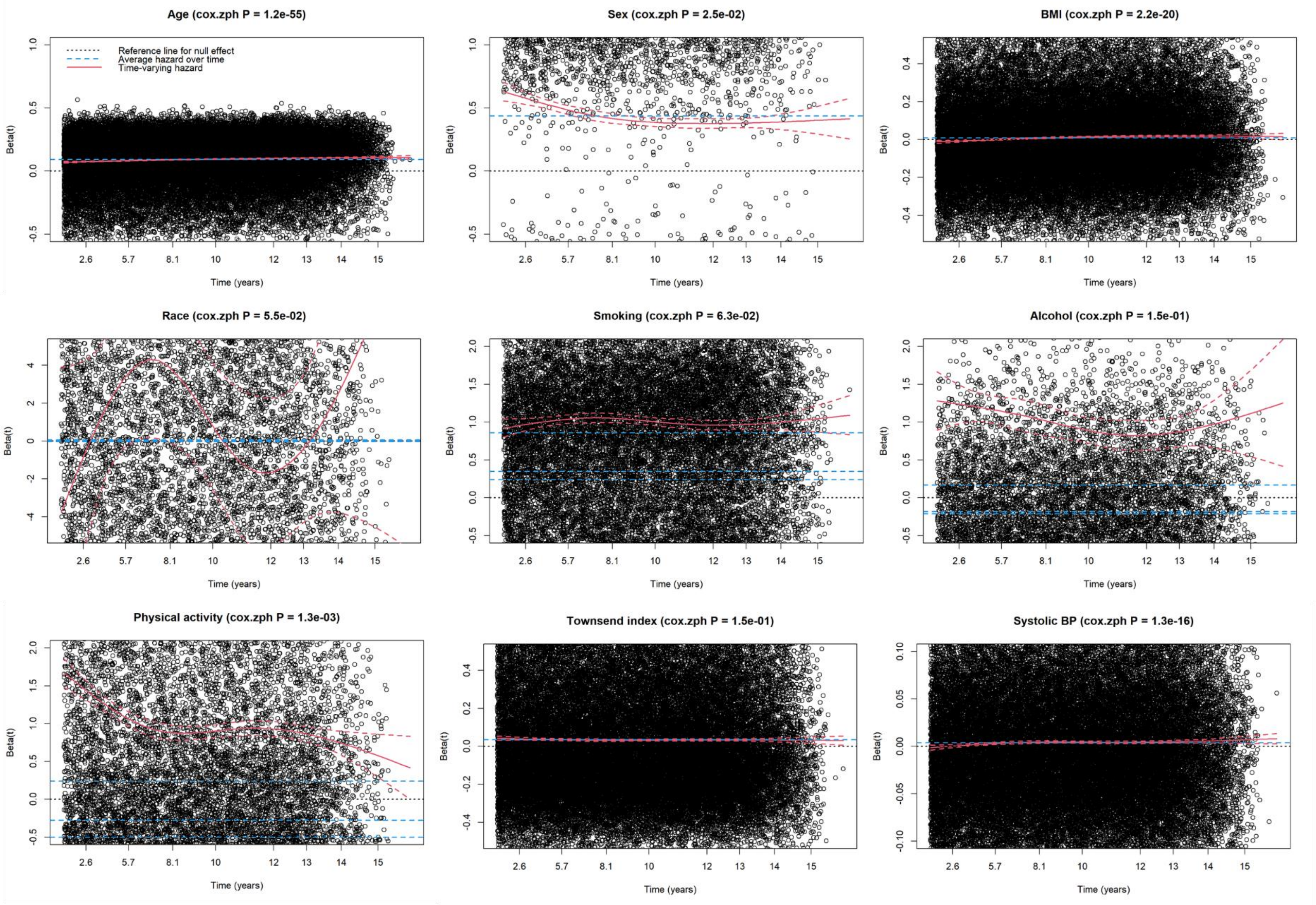
Age (cox.zph P = 1.2e-55)
Reference line for null effect
Average hazard over time
Time-varying hazard
Sex (cox.zph P = 2.5e-02)
BMI (cox.zph P = 2.2e-20)
Race (cox.zph P = 5.5e-02)
Smoking (cox.zph P = 6.3e-02)
Alcohol (cox.zph P = 1.5e-01)
Physical activity (cox.zph P = 1.3e-03)
Townsend index (cox.zph P = 1.5e-01)
Systolic BP (cox.zph P = 1.3e-16)
Beta(t)
Time (years)
2.6
5.7
8.1
10
12
13
14
15

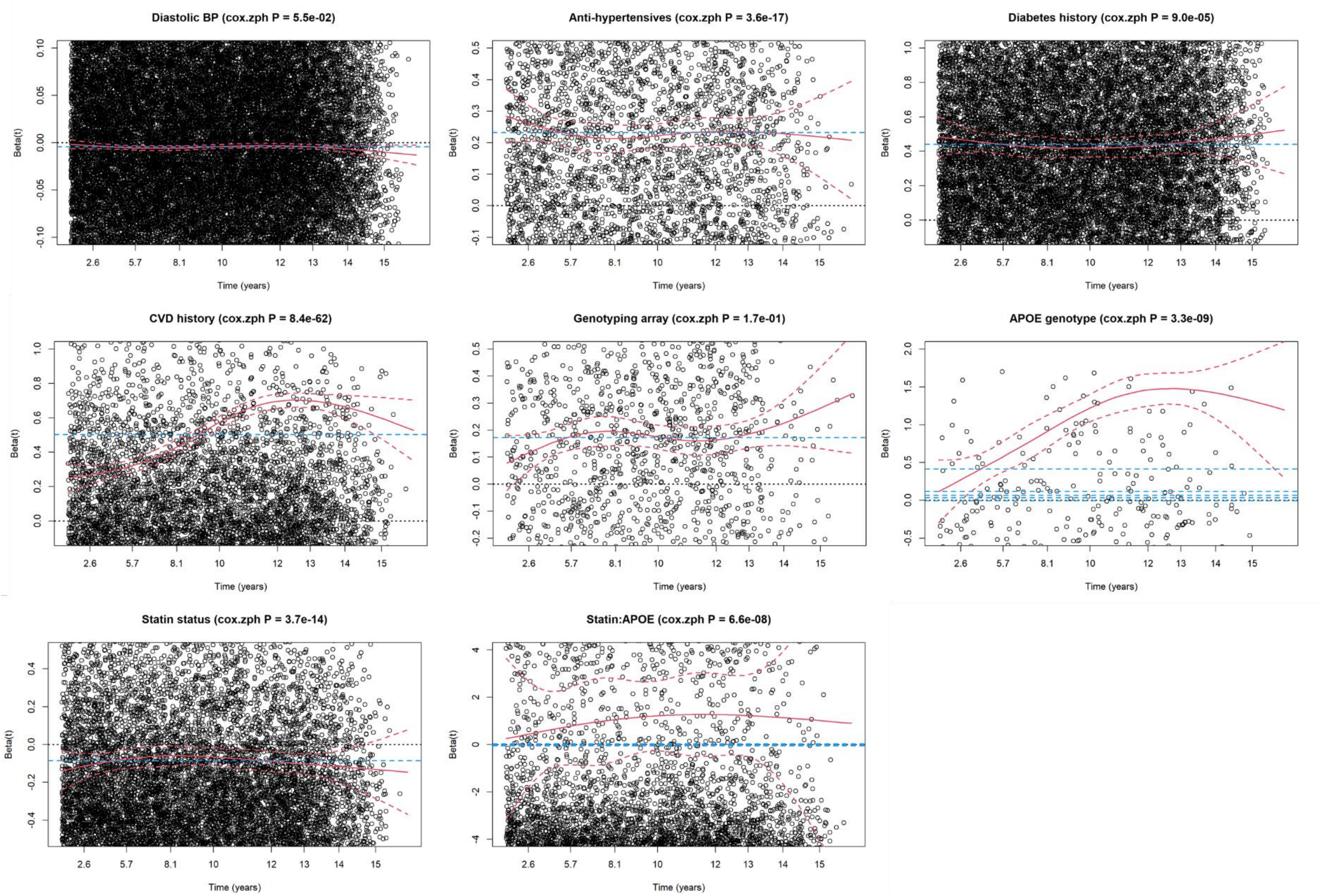


**Figure S3. Results of the proportional hazards assumption test for the analysis of all-cause death in the UK Biobank Baseline Analysis.** *APOE* = Apolipoprotein E, BMI = body mass index, CVD = cardiovascular disease.
